# Sex disparities in outcome of medication-assisted therapy of opioid use disorder: Nationally representative study

**DOI:** 10.1101/2024.09.24.24314320

**Authors:** Eduardo R. Butelman, Yuefeng Huang, Alicia McFarlane, Carolann Slattery, Rita Z. Goldstein, Nora D. Volkow, Nelly Alia-Klein

## Abstract

**Question:** The opioid epidemic causes massive morbidity, and males have substantially greater overdose mortality rates than females. It is unclear whether there are sex-related disparities at different stages in the trajectory of opioid use disorders, in “real world” settings.

**Goal:** To determine sex disparities in non-medical opioid use (NMOU) at the end of outpatient medication-assisted treatment (MAT), using nationally representative data.

**Design:** Observational epidemiological study of publicly funded outpatient MAT programs in the national “Treatment episode data set-discharges” (TEDS-D) for 2019.

**Participants:** Persons aged ≥18 in their first treatment episode, in outpatient MAT for use of heroin or other opioids (N=11,549). The binary outcome was presence/absence of NMOU.

**Results:** In univariate analyses, males had significantly higher odds of NMOU, compared to females (odds ratio=1.27; Chi^2^ [df:1]=39.08; uncorrected p<0.0001; p=0.0041 after Bonferroni correction). A multivariable logistic regression detected a male>female odds ratio of 1.19 (95%CI=1.09-1.29; p<0.0001), adjusting for socio-demographic/clinical variables. Several specific conditions were revealed in which males had greater odds of NMOU compared to females (e.g., at ages 18-29 and 30-39; corrected p=0.012, or if they used opioids by inhalation; corrected p=0.0041).

**Conclusions:** This nationally representative study indicates that males have greater odds of NMOU in their first episode of MAT, indicating more unfavorable outcomes. The study reveals specific socio-demographic and clinical variables under which this sex disparity is most prominent.

**Highlights:** **\***It is unclear if there are sex-related disparities in outcomes for outpatient opioid medication-assisted therapy (MAT), in large-scale “real world” settings.

**\***In this nationally representative “real world” study, adult males had significantly greater odds of non-medical opioid use (NMOU) in the month prior to discharge from their first MAT episode compared to females, adjusting for socio-demographic and clinical variables. Males were at higher risk than females for this undesirable outcome under several conditions (e.g., in younger age categories, or if their route of NMOU was by inhalation.

**\***Sex disparities in MAT outcomes occur under specific conditions that can be examined and potentially addressed, with the goal of improving personalized approaches for OUD.

## 1. Introduction

The opioid epidemic causes massive health consequences in the U.S. (Butelman et al., 2023; Wilson et al., 2020), and we recently reported approximately 2.5-fold greater opioid-induced overdose mortality rates in males versus females, even after adjusting for sex-specific frequency of opioid use (Butelman et al., 2023). Therefore, this sex disparity was not simply due to differing levels of overall opioid use, and may reflect multiple biological, socio-demographic and clinical factors, as well as response to treatment (Apsley et al., 2024; Little and Kosten, 2023; McEvoy et al., 2023; Towers et al., 2023). Understanding these sex differences could inform the development of personalized sex or gender-specific interventions, and decrease morbidity and mortality (Apsley et al., 2024; Kreek et al., 2019; Marsh et al., 2021; Strang et al., 2020).

Research studies have compared different stages in the trajectory of opioid use disorders (OUD) in males and females, as well as response to medication-assisted treatment (MAT) (Bawor et al., 2015; Marsh et al., 2021; Zheng et al., 2024). However, it is difficult to draw overall conclusions across studies on MAT outcomes, based on differences across time, cohort, study methodology and environment (Bawor et al., 2015; Huhn et al., 2019; Zheng et al., 2024).

It has been noted that some research studies may not be representative of “real world” MAT settings, due to relative lack of inclusion based on socio-demographic factors, which can also be related to social determinants of health (Braveman and Gottlieb, 2014; Eghaneyan et al., 2020; Rudolph et al., 2022). Intriguingly, a “real world” study (using the 2017 Treatment Episode Data Set-Discharges; TEDS-D) detected that females in MAT had 1.1 greater odds of staying in treatment at least 6 months, compared to males (Krawczyk et al., 2021). However, duration of treatment in MAT can differ because of environmental and program-level features, in addition to client-level factors (Durand et al., 2021; Krawczyk et al., 2021). Other outcomes such as “treatment completion” may also be influenced by program-level features, racial/ethnic disparities, and socioeconomic disadvantages and social determinants of health (Askari et al., 2020; Franz et al., 2023; Mennis et al., 2019). A major favorable clinical outcome of MAT is decreased NMOU (Goldstein, 2022; Kreek et al., 2019; Strang et al., 2020). Therefore, this study examined whether there are sex differences in the frequency of NMOU in the month prior to discharge from MAT (Burgess-Hull et al., 2022; Kowalczyk et al., 2015), in a nationally representative “real world” outpatient sample, adjusting for length of stay in treatment, and major socio-demographic and clinical variables (Fischer et al., 2023; Stahler and Mennis, 2018; Surratt et al., 2011).

## 2. Methods

### 2.1 Data Source

This study used the Treatment Episode Data Set-Discharges (TEDS-D), from 2019 (https://www.datafiles.samhsa.gov/), the newest data available prior to the disruptions of the COVID-19 pandemic (Volkow, 2020). The TEDS-D is composed of individual episodes from substance use treatment facilities that receive public funding; data includes the continental US, excluding Oregon, Washington state and West Virginia. Data extraction and variable coding are described below.

### 2.2 Outcome variable

The outcome variable was ***NMOU in the month prior to discharge*** (TEDS-D variable FREQ1_D), recoded as a binary variable, combining “some use” and “daily use”, versus “no use”.

### 2.3 Inclusion Criteria

**A)** <u>First episodes of treatment</u> (variable NOPRIOR=0). This criterion avoids confounding due to potentially examining several episodes in the same person, or due to prior treatment history (Huhn et al., 2018; Kitsantas et al., 2023). **B)** Episodes in which an <u>opioid is the primary substance</u> (i.e., heroin, non-prescription methadone, or “other opiates and synthetics”) at admission and discharge (i.e., variables SUB1 and SUB1_D=5,6, or 7, respectively). **C)** Episodes with MAT in the treatment plan (variable METHUSE=1; principally methadone and buprenorphine, but also naltrexone) (Ross et al., 2024; Strang et al., 2020; Wakeman et al., 2020). The “METHUSE” variable pools these medications, not allowing further stratification. **D)** Service in an <u>ambulatory (outpatient) setting</u> (variable SERVICES=6 or 7) (Kreek et al., 2019; Strang et al., 2020; Wakeman et al., 2020). **E)** Episodes with a <u>length of stay</u> (variable LOS) of ≥30 days, to focus on those who had at least initial treatment engagement (Blanco and Volkow, 2019; Krawczyk et al., 2021). **F)** Episodes in <u>census divisions</u> 1-9 (i.e., continental U.S).

### 2.4 Exclusion criteria

**A)** <u>Age <18</u>, due to low prevalence. **B)** <u>Episodes terminated because the person had died</u> (cause of death is unavailable; variable REASON=5), or became <u>incarcerated</u> (variable REASON=6). **C)** Episodes in which the route of non-medical opioid use was “other” (i.e., variable ROUTE1=5), due to low prevalence. **D) Missing data:** If there were missing data in any of these variables, the episode was excluded from analysis (episode-wise deletion).

### 2.5 Demographic variables

In addition to sex, several variables were examined: Length of stay in treatment (variable LOS), coded in bins denoting increasing duration (31-45, 46-60, 61-90, 91-180, 181-365, ≥366 days). Route of NMOU (variable ROUTE1) was coded as four categories: oral, smoking, inhalation or injection. Age at admission was recoded as 18-29, 30-39, 40-49, ≥50 age bins. Housing at discharge was coded as: independent housing, dependent (e.g., supportive) housing, and homeless (unhoused). Employment at discharge was coded as: employed full time, employed part-time, “not in the labor force” and unemployed. Racial background was studied in three categories (African-American/black, white, and all remaining recoded as “other”). Psychiatric comorbidity was coded as a binary variable (present/absent). Hispanic/latino ethnicity was re-coded as a binary variable (any hispanic/latino versus non-hispanic/latino). Referral source was re-coded as a binary variable, with court/criminal justice referrals versus all others. Census divisions were also examined.

### 2.6 Data analyses

Analyses were carried out with GraphPad Prism (V. 10) and TIBCO Statistica Data Science Workbench. The alpha-level of significance was p=0.05.

#### 2.6.1 Univariate Analyses

Demographics were examined with Chi^2^ analyses across sex. The relative frequency of the outcome (use vs no use of non-medical opioids) in males and females was also examined in Chi^2^ 2X2 contingency analyses. The p-values for these contingency analyses were Bonferroni-corrected, and male/female odds ratios are shown for those that survive correction. 95%CI for the proportions of males and females with the outcome were calculated with the Wilson-Brown method (Brown et al., 2001). Data are also presented as % of males and females with non-medical opioid use, and male-female difference.

#### 2.6.2 Multivariable analysis

A multivariable logistic regression (Hosmer and Lemeshow, 2004; Osborne, 2014) using maximum likelihood estimation was carried out for the binary outcome of NMOU, versus no use. Sex and all the variables above were entered in the regression. Adjusted Beta-parameters and odds ratios of the outcome are reported. Regression performance was examined with receiver operating characteristic (ROC) analysis, Nagelkerke R^2^ and likelihood ratio test (Meurer and Tolles, 2017).

### 2.7 Sensitivity Analyses

**a)** Exploration of interaction terms between sex and other relevant variables in the multivariable regression model, **b)** Examination of the subset whose primary drug was heroin (as opposed the combined group with heroin, non-medical methadone and “other opiates and synthetics”), and **c)** Examination of major poly-drug use patterns, with the subset who had an opioid as primary substance *and* either cocaine or methamphetamine as a secondary substance (using variable SUB2).

## 3. Results

After the inclusion and exclusion criteria above were satisfied, and deletion of episodes with missing data in any of the studied variables, n=11,549 episodes available for analysis (see Supplement Flowchart).

### 3.1 Sample demographics by sex

The number of males and females for each condition is in Supplementary Table S1, examined with Chi^2^ contingency analyses. Among sex disparities observed, there was a smaller relative frequency of males versus females with a psychiatric comorbidity.

#### 3.2.1 Univariate association of sex with the outcome (NMOU)

Overall, 61.8% of males and 56.0% of females had NMOU. The 2X2 contingency analysis (male/female X use/no use) was significant (Chi^2^[1]=39.08; uncorrected p<0.0001; Bonferroni-corrected p=0.0041) with a male/female unadjusted odds ratio of =1.27 (95%CI: 1.18-1.38) (Table 1; Figure 1).

**Table 1:**
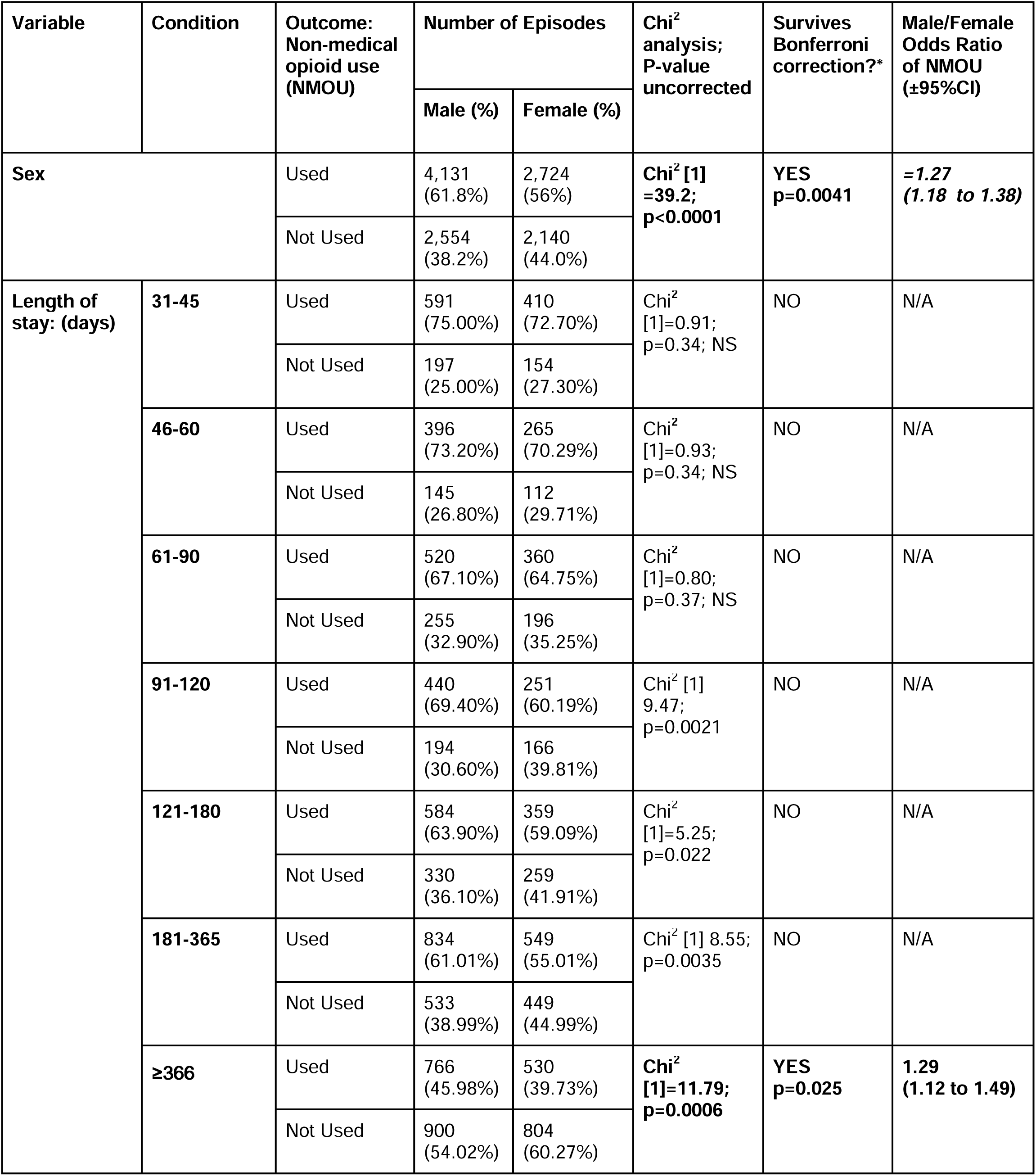
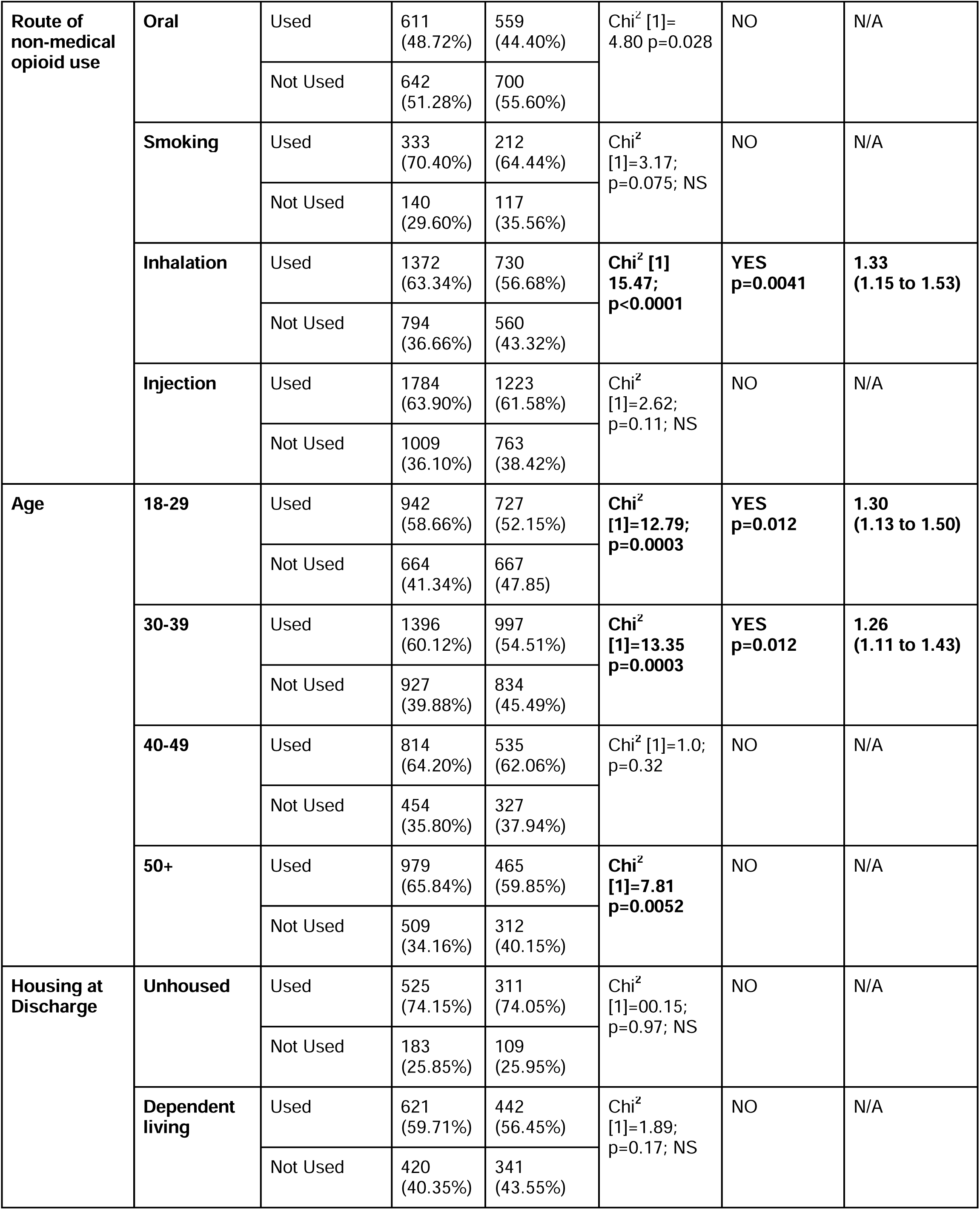
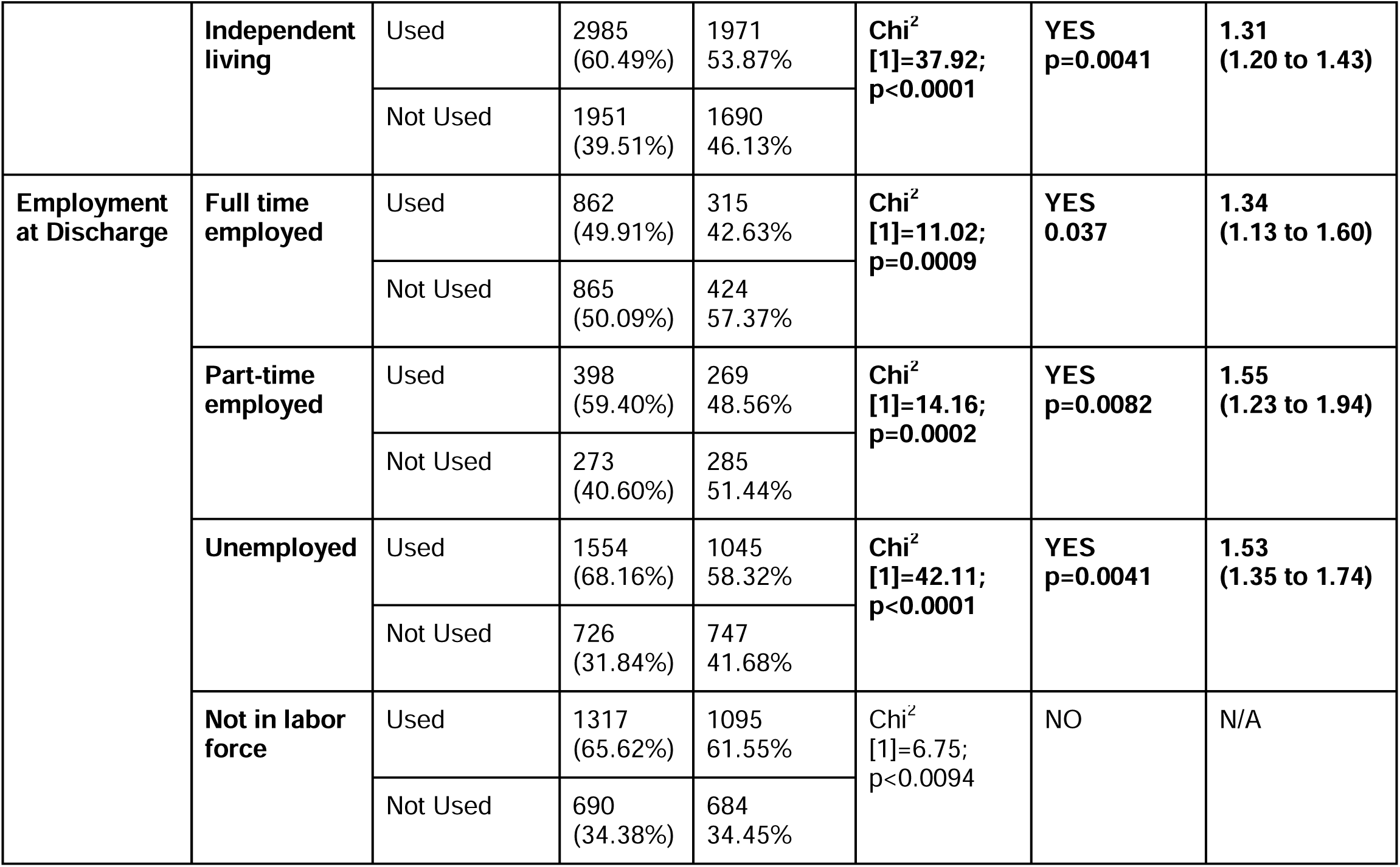
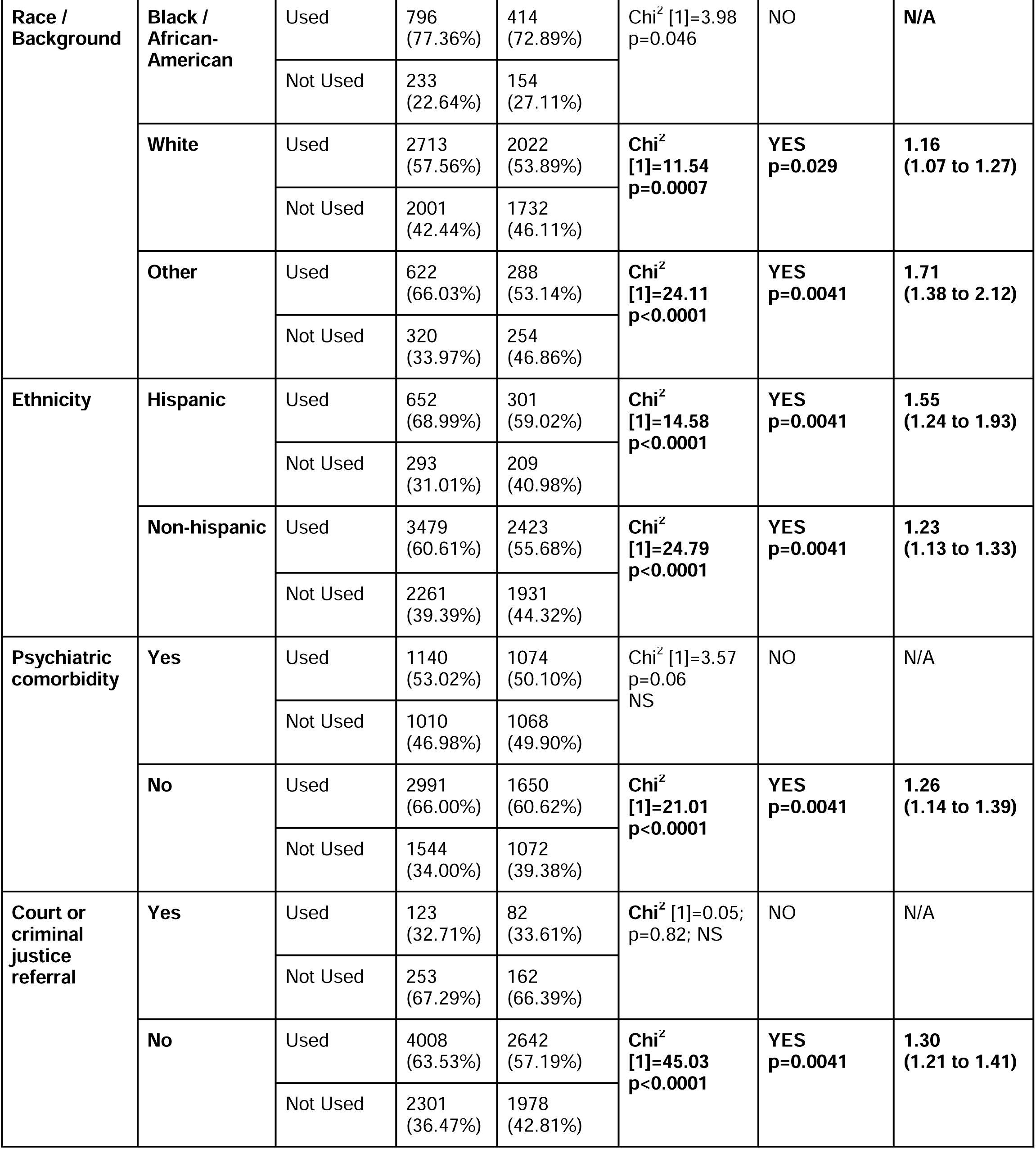
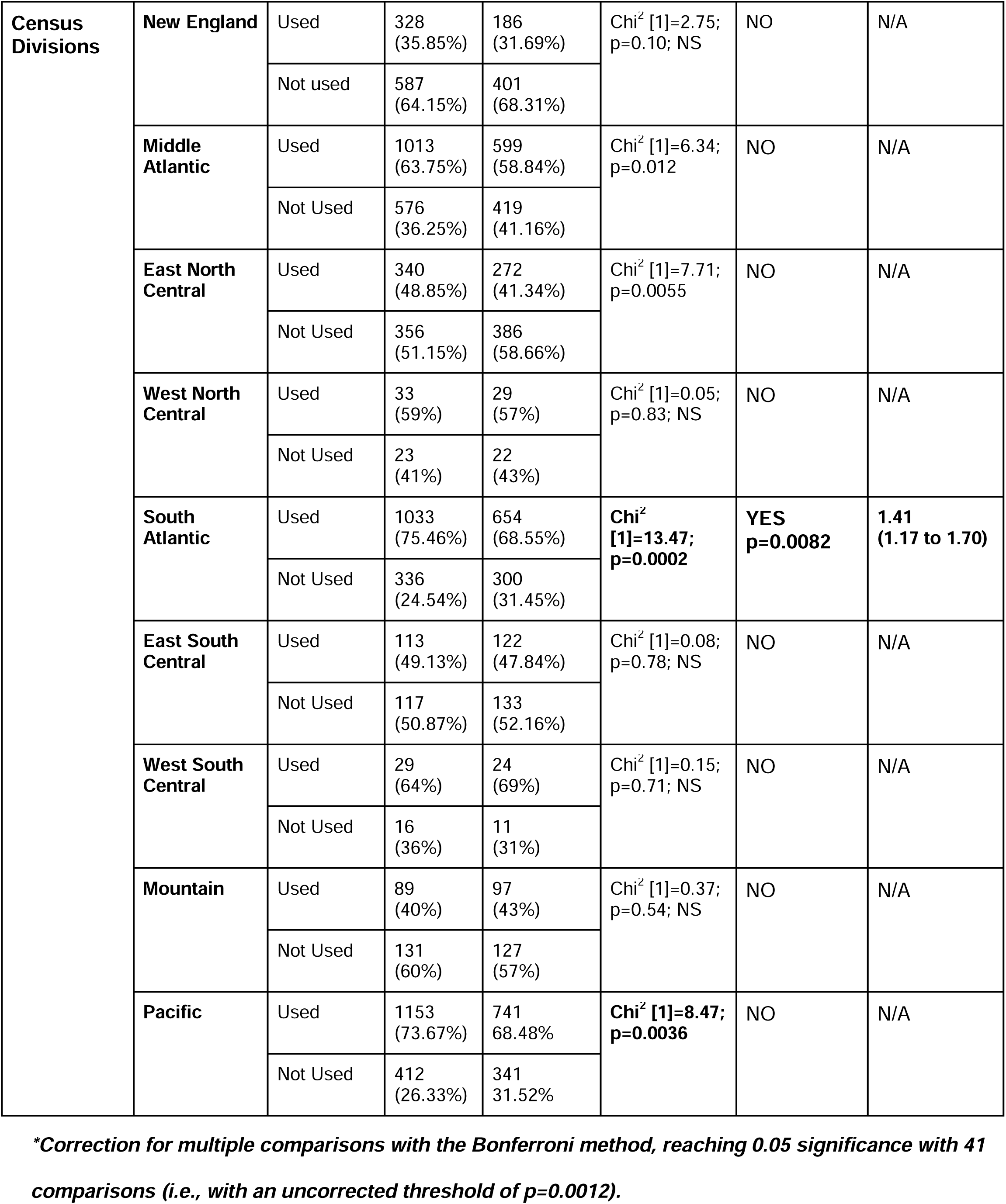
Sex-related association with the outcome: NMOU in the month prior to discharge (2X2 contingency analyses; univariate)

| Variable | Condition | Outcome:<br>Non-medical<br>opioid use<br>(NMOU) | Number of Episodes |  | Chi<sup>2</sup><br>analysis;<br>P-value<br>uncorrected | Survives<br>Bonferroni<br>correction?* | Male/Female<br>Odds Ratio<br>of NMOU<br>(±95%CI) |
| --- | --- | --- | --- | --- | --- | --- | --- |
|  |  |  | Male (%) | Female (%) |  |  |  |
| **Sex** |  | Used | 4,131<br>(61.8%) | 2,724<br>(56%) | **Chi<sup>2</sup> [1]**<br>**=39.2;**<br>**p<0.0001** | **YES**<br>**p=0.0041** | **=1.27**<br>**(1.18 to 1.38)** |
|  |  | Not Used | 2,554<br>(38.2%) | 2,140<br>(44.0%) |  |  |  |
| **Length of**<br>**stay: (days)** | **31-45** | Used | 591<br>(75.00%) | 410<br>(72.70%) | Chi<sup>2</sup><br>[1]=0.91;<br>p=0.34; NS | NO | N/A |
|  |  | Not Used | 197<br>(25.00%) | 154<br>(27.30%) |  |  |  |
|  | **46-60** | Used | 396<br>(73.20%) | 265<br>(70.29%) | Chi<sup>2</sup><br>[1]=0.93;<br>p=0.34; NS | NO | N/A |
|  |  | Not Used | 145<br>(26.80%) | 112<br>(29.71%) |  |  |  |
|  | **61-90** | Used | 520<br>(67.10%) | 360<br>(64.75%) | Chi<sup>2</sup><br>[1]=0.80;<br>p=0.37; NS | NO | N/A |
|  |  | Not Used | 255<br>(32.90%) | 196<br>(35.25%) |  |  |  |
|  | **91-120** | Used | 440<br>(69.40%) | 251<br>(60.19%) | Chi<sup>2</sup> [1]<br>9.47;<br>p=0.0021 | NO | N/A |
|  |  | Not Used | 194<br>(30.60%) | 166<br>(39.81%) |  |  |  |
|  | **121-180** | Used | 584<br>(63.90%) | 359<br>(59.09%) | Chi<sup>2</sup><br>[1]=5.25;<br>p=0.022 | NO | N/A |
|  |  | Not Used | 330<br>(36.10%) | 259<br>(41.91%) |  |  |  |
|  | **181-365** | Used | 834<br>(61.01%) | 549<br>(55.01%) | Chi<sup>2</sup> [1] 8.55;<br>p=0.0035 | NO | N/A |
|  |  | Not Used | 533<br>(38.99%) | 449<br>(44.99%) |  |  |  |
|  | **≥366** | Used | 766<br>(45.98%) | 530<br>(39.73%) | **Chi<sup>2</sup>**<br>**[1]=11.79;**<br>**p=0.0006** | **YES**<br>**p=0.025** | **1.29**<br>**(1.12 to 1.49)** |
|  |  | Not Used | 900<br>(54.02%) | 804<br>(60.27%) |  |  |  |
| **Route of non-medical opioid use** | **Oral** | Used | 611<br>(48.72%) | 559<br>(44.40%) | Chi<sup>2</sup> [1]=<br>4.80 p=0.028 | NO | N/A |
|  |  | Not Used | 642<br>(51.28%) | 700<br>(55.60%) |  |  |  |
|  | **Smoking** | Used | 333<br>(70.40%) | 212<br>(64.44%) | Chi<sup>2</sup><br>[1]=3.17;<br>p=0.075; NS | NO | N/A |
|  |  | Not Used | 140<br>(29.60%) | 117<br>(35.56%) |  |  |  |
|  | **Inhalation** | Used | 1372<br>(63.34%) | 730<br>(56.68%) | **Chi<sup>2</sup> [1]**<br>**15.47;**<br>**p<0.0001** | **YES**<br>**p=0.0041** | **1.33**<br>**(1.15 to 1.53)** |
|  |  | Not Used | 794<br>(36.66%) | 560<br>(43.32%) |  |  |  |
|  | **Injection** | Used | 1784<br>(63.90%) | 1223<br>(61.58%) | Chi<sup>2</sup><br>[1]=2.62;<br>p=0.11; NS | NO | N/A |
|  |  | Not Used | 1009<br>(36.10%) | 763<br>(38.42%) |  |  |  |
| **Age** | **18-29** | Used | 942<br>(58.66%) | 727<br>(52.15%) | **Chi<sup>2</sup>**<br>**[1]=12.79;**<br>**p=0.0003** | **YES**<br>**p=0.012** | **1.30**<br>**(1.13 to 1.50)** |
|  |  | Not Used | 664<br>(41.34%) | 667<br>(47.85%) |  |  |  |
|  | **30-39** | Used | 1396<br>(60.12%) | 997<br>(54.51%) | **Chi<sup>2</sup>**<br>**[1]=13.35**<br>**p=0.0003** | **YES**<br>**p=0.012** | **1.26**<br>**(1.11 to 1.43)** |
|  |  | Not Used | 927<br>(39.88%) | 834<br>(45.49%) |  |  |  |
|  | **40-49** | Used | 814<br>(64.20%) | 535<br>(62.06%) | Chi<sup>2</sup> [1]=1.0;<br>p=0.32 | NO | N/A |
|  |  | Not Used | 454<br>(35.80%) | 327<br>(37.94%) |  |  |  |
|  | **50+** | Used | 979<br>(65.84%) | 465<br>(59.85%) | **Chi<sup>2</sup>**<br>**[1]=7.81**<br>**p=0.0052** | NO | N/A |
|  |  | Not Used | 509<br>(34.16%) | 312<br>(40.15%) |  |  |  |
| **Housing at Discharge** | **Unhoused** | Used | 525<br>(74.15%) | 311<br>(74.05%) | Chi<sup>2</sup><br>[1]=00.15;<br>p=0.97; NS | NO | N/A |
|  |  | Not Used | 183<br>(25.85%) | 109<br>(25.95%) |  |  |  |
|  | **Dependent living** | Used | 621<br>(59.71%) | 442<br>(56.45%) | Chi<sup>2</sup><br>[1]=1.89;<br>p=0.17; NS | NO | N/A |
|  |  | Not Used | 420<br>(40.35%) | 341<br>(43.55%) |  |  |  |
|  | **Independent living** | Used | 2985<br>(60.49%) | 1971<br>53.87% | **Chi<sup>2</sup>**<br>**[1]=37.92;**<br>**p<0.0001** | **YES**<br>**p=0.0041** | **1.31**<br>**(1.20 to 1.43)** |
|  |  | Not Used | 1951<br>(39.51%) | 1690<br>46.13% |  |  |  |
| **Employment at Discharge** | **Full time employed** | Used | 862<br>(49.91%) | 315<br>42.63% | **Chi<sup>2</sup>**<br>**[1]=11.02;**<br>**p=0.0009** | **YES**<br>**0.037** | **1.34**<br>**(1.13 to 1.60)** |
|  |  | Not Used | 865<br>(50.09%) | 424<br>57.37% |  |  |  |
|  | **Part-time employed** | Used | 398<br>(59.40%) | 269<br>48.56% | **Chi<sup>2</sup>**<br>**[1]=14.16;**<br>**p=0.0002** | **YES**<br>**p=0.0082** | **1.55**<br>**(1.23 to 1.94)** |
|  |  | Not Used | 273<br>(40.60%) | 285<br>51.44% |  |  |  |
|  | **Unemployed** | Used | 1554<br>(68.16%) | 1045<br>58.32% | **Chi<sup>2</sup>**<br>**[1]=42.11;**<br>**p<0.0001** | **YES**<br>**p=0.0041** | **1.53**<br>**(1.35 to 1.74)** |
|  |  | Not Used | 726<br>(31.84%) | 747<br>41.68% |  |  |  |
|  | **Not in labor force** | Used | 1317<br>(65.62%) | 1095<br>61.55% | **Chi<sup>2</sup>**<br>**[1]=6.75;**<br>**p<0.0094** | **NO** | **N/A** |
|  |  | Not Used | 690<br>(34.38%) | 684<br>34.45% |  |  |  |

| Table 1<br>Continued | Condition | Outcome | Number of Episodes |  | Chi-square<br>analysis;<br>P-value | Survives<br>correction?* | Male/Female<br>Odds Ratio<br>of non-medical<br>opioid use<br>(±95%CI) |
| --- | --- | --- | --- | --- | --- | --- | --- |
|  |  |  | Male (%) | Female (%) |  |  |  |
| Race /<br>Background | Black /<br>African-American | Used | 796<br>(77.36%) | 414<br>(72.89%) | Chi<sup>2</sup> [1]=3.98<br>p=0.046 | NO | N/A |
|  |  | Not Used | 233<br>(22.64%) | 154<br>(27.11%) |  |  |  |
|  | White | Used | 2713<br>(57.56%) | 2022<br>(53.89%) | Chi<sup>2</sup><br>[1]=11.54<br>p=0.0007 | YES<br>p=0.029 | 1.16<br>(1.07 to 1.27) |
|  |  | Not Used | 2001<br>(42.44%) | 1732<br>(46.11%) |  |  |  |
|  | Other | Used | 622<br>(66.03%) | 288<br>(53.14%) | Chi<sup>2</sup><br>[1]=24.11<br>p<0.0001 | YES<br>p=0.0041 | 1.71<br>(1.38 to 2.12) |
|  |  | Not Used | 320<br>(33.97%) | 254<br>(46.86%) |  |  |  |
| Ethnicity | Hispanic | Used | 652<br>(68.99%) | 301<br>(59.02%) | Chi<sup>2</sup><br>[1]=14.58<br>p<0.0001 | YES<br>p=0.0041 | 1.55<br>(1.24 to 1.93) |
|  |  | Not Used | 293<br>(31.01%) | 209<br>(40.98%) |  |  |  |
|  | Non-hispanic | Used | 3479<br>(60.61%) | 2423<br>(55.68%) | Chi<sup>2</sup><br>[1]=24.79<br>p<0.0001 | YES<br>p=0.0041 | 1.23<br>(1.13 to 1.33) |
|  |  | Not Used | 2261<br>(39.39%) | 1931<br>(44.32%) |  |  |  |
| Psychiatric<br>comorbidity | Yes | Used | 1140<br>(53.02%) | 1074<br>(50.10%) | Chi<sup>2</sup> [1]=3.57<br>p=0.06<br>NS | NO | N/A |
|  |  | Not Used | 1010<br>(46.98%) | 1068<br>(49.90%) |  |  |  |
|  | No | Used | 2991<br>(66.00%) | 1650<br>(60.62%) | Chi<sup>2</sup><br>[1]=21.01<br>p<0.0001 | YES<br>p=0.0041 | 1.26<br>(1.14 to 1.39) |
|  |  | Not Used | 1544<br>(34.00%) | 1072<br>(39.38%) |  |  |  |
| Court or<br>criminal<br>justice<br>referral | Yes | Used | 123<br>(32.71%) | 82<br>(33.61%) | Chi<sup>2</sup> [1]=0.05;<br>p=0.82; NS | NO | N/A |
|  |  | Not Used | 253<br>(67.29%) | 162<br>(66.39%) |  |  |  |
|  | No | Used | 4008<br>(63.53%) | 2642<br>(57.19%) | Chi<sup>2</sup><br>[1]=45.03<br>p<0.0001 | YES<br>p=0.0041 | 1.30<br>(1.21 to 1.41) |
|  |  | Not Used | 2301<br>(36.47%) | 1978<br>(42.81%) |  |  |  |
| **Census Divisions** | **New England** | Used | 328<br>(35.85%) | 186<br>(31.69%) | Chi<sup>2</sup> [1]=2.75;<br>p=0.10; NS | NO | N/A |
|  |  | Not used | 587<br>(64.15%) | 401<br>(68.31%) |  |  |  |
|  | **Middle Atlantic** | Used | 1013<br>(63.75%) | 599<br>(58.84%) | Chi<sup>2</sup> [1]=6.34;<br>p=0.012 | NO | N/A |
|  |  | Not Used | 576<br>(36.25%) | 419<br>(41.16%) |  |  |  |
|  | **East North Central** | Used | 340<br>(48.85%) | 272<br>(41.34%) | Chi<sup>2</sup> [1]=7.71;<br>p=0.0055 | NO | N/A |
|  |  | Not Used | 356<br>(51.15%) | 386<br>(58.66%) |  |  |  |
|  | **West North Central** | Used | 33<br>(59%) | 29<br>(57%) | Chi<sup>2</sup> [1]=0.05;<br>p=0.83; NS | NO | N/A |
|  |  | Not Used | 23<br>(41%) | 22<br>(43%) |  |  |  |
|  | **South Atlantic** | Used | 1033<br>(75.46%) | 654<br>(68.55%) | **Chi<sup>2</sup> [1]=13.47;**<br>**p=0.0002** | **YES**<br>**p=0.0082** | **1.41**<br>**(1.17 to 1.70)** |
|  |  | Not Used | 336<br>(24.54%) | 300<br>(31.45%) |  |  |  |
|  | **East South Central** | Used | 113<br>(49.13%) | 122<br>(47.84%) | Chi<sup>2</sup> [1]=0.08;<br>p=0.78; NS | NO | N/A |
|  |  | Not Used | 117<br>(50.87%) | 133<br>(52.16%) |  |  |  |
|  | **West South Central** | Used | 29<br>(64%) | 24<br>(69%) | Chi<sup>2</sup> [1]=0.15;<br>p=0.71; NS | NO | N/A |
|  |  | Not Used | 16<br>(36%) | 11<br>(31%) |  |  |  |
|  | **Mountain** | Used | 89<br>(40%) | 97<br>(43%) | Chi<sup>2</sup> [1]=0.37;<br>p=0.54; NS | NO | N/A |
|  |  | Not Used | 131<br>(60%) | 127<br>(57%) |  |  |  |
|  | **Pacific** | Used | 1153<br>(73.67%) | 741<br>68.48% | **Chi<sup>2</sup> [1]=8.47;**<br>**p=0.0036** | NO | N/A |
|  |  | Not Used | 412<br>(26.33%) | 341<br>31.52% |  |  |  |

***\*Correction for multiple comparisons with the Bonferroni method, reaching 0.05 significance with 41 comparisons (i.e., with an uncorrected threshold of p=0.0012).***

**Fig. 1A:**
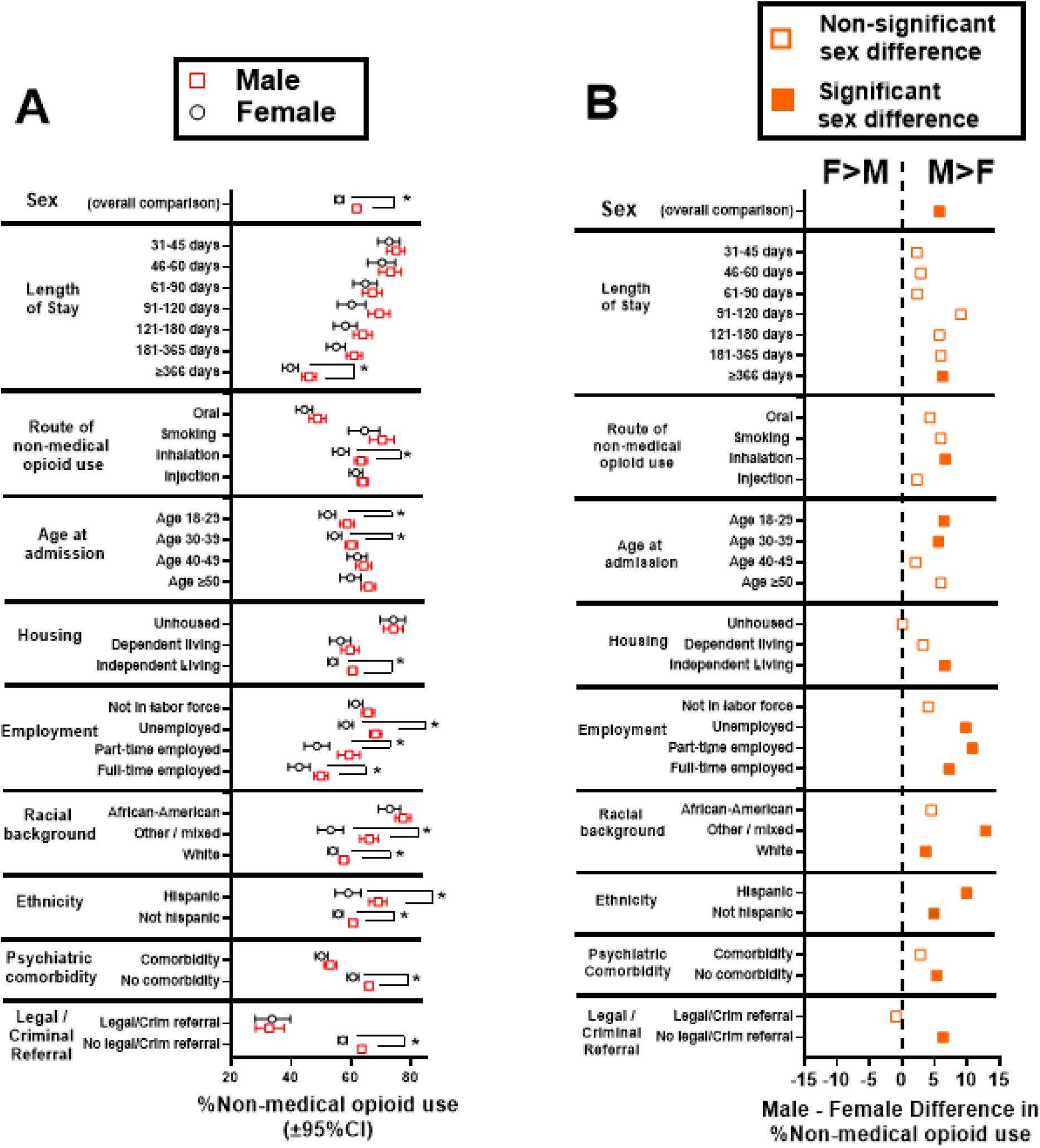
Percent of males and females with NMOU; **\***Conditions with significant 2X2 contingency analyses by sex, after Bonferroni correction (see Table 1, which includes data for census divisions). **Fig 1B**: Male-Female differences in %NMOU (data from Fig. 1A). Positive values indicate greater %NMOU in males vs females (labeled **“M>F”** at the top), and negative values indicate greater %NMOU in females vs males (labeled **“F>M”**). Filled symbols indicate sex comparisons that survive Bonferroni corrections (Table 1); open symbols indicate comparisons that did not reach significance.

#### 3.2.2 Univariate association of demographic and clinical variables with NMOU, by sex

Contingency analyses examined sex-associated outcomes (Table 1). Figure 1A shows %non-medical opioid use in males and females; Figure 1B shows the Male-Female difference. After correction for multiple comparisons, males had greater odds of NMOU compared to females in the following conditions: **a)** at the longest lengths of stay (i.e., ≥366 days), **b)** if NMOU was by inhalation, **c)** in the younger age bins (i.e., 18-29 and 30-39), **d)** in independent housing, **e)** employed full-time, part-time or unemployed, **f)** white or “mixed” racial categories **g)** whether they were of hispanic/latino ethnicity or not, **h)** if they did not have a psychiatric comorbidity, **i)** if they did not have a court/criminal referral to treatment and **j)** in the South Atlantic census division. Importantly, females did not have greater odds of NMOU than males in any of the conditions under study, after multiple comparison correction (Figure 1A and 1B).

#### 3.2.3 Multivariable logistic regression of NMOU as the outcome

Adjusted odds ratios of NMOU are in Figure 2 and Supplement Table S2 (which also includes data for census divisions). **Overall regression performance:** The regression had an area under the receiver operating curve (AUROC)=73.8, Nagelkerke R^2^=0.22 and likelihood ratio G^2^=2,027 [df=31; p<0.0001).

**Fig. 2:**
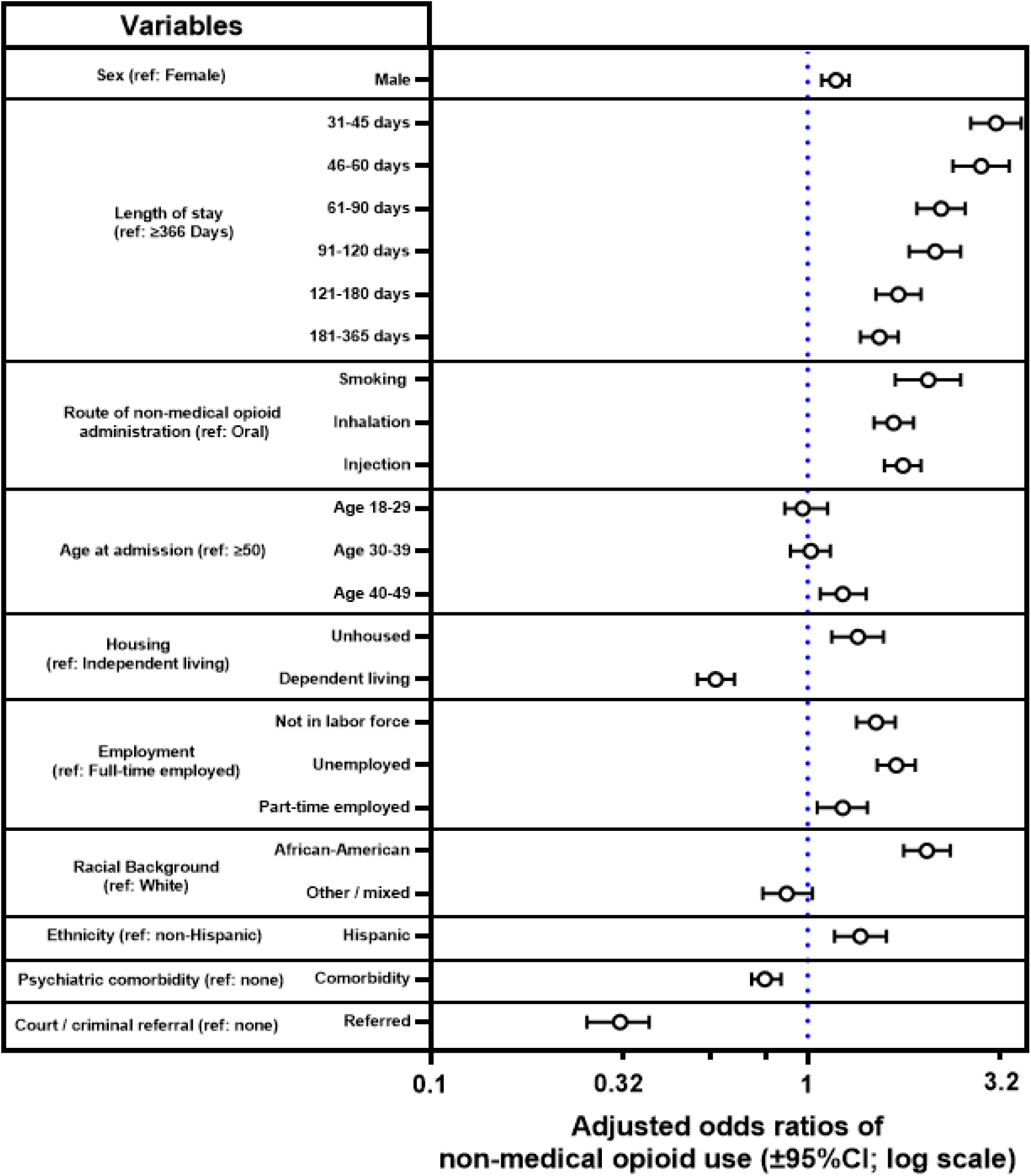
Adjusted odds ratios of NMOU, from multivariable logistic regression. Label “ref.” indicates the reference category for each variable. Data for census divisions and full regression parameters are in the Supplement (Table S2).

#### Sex

Males had an adjusted odds ratio =1.19 (95%CI: 1.089-1.291; p<0.0001) of NMOU, compared to females. **Length of stay in treatment:** All of the lengths of stay <366 days were associated with greater odds of NMOU, compared to the reference category (≥366 days). The greatest odds ratios were observed at the shorter lengths of stay (e.g., 31-45 and 46-60 days). **Route of NMOU:** Those with inhalation, smoking, or injection as route of administration had greater odds of NMOU, compared to the oral route. **Age:** Persons in the age range 40-49 had increased odds of NMOU, compared to the reference category (≥50). **Housing:** Persons who were unhoused had increased odds of NMOU, compared to those in independent living, whereas those in “dependent” living (e.g., supportive housing) had decreased odds. **Employment:** Those who were unemployed or not in the labor force had greater odds of NMOU, compared to being employed full-time. **Racial background:** African-American/black persons had greater odds of NMOU, compared to white persons. **Ethnicity:** Persons of hispanic/latino ethnicity also had increased odds of NMOU, compared to non-hispanic/latinos. **Psychiatric comorbidity:** Persons with psychiatric comorbidities had lower odds of NMOU, compared to those without such comorbidity. **Court or criminal referral to treatment:** Persons entering treatment through court/criminal referral had lower odds, compared to those without such a referral. **Census divisions:** Compared to the reference category (Middle Atlantic), those in the Pacific and South Atlantic divisions had greater odds of NMOU, whereas those in New England, East North Central, East South Central and Mountain had lower odds (see Table S2).

### 3.3 Sensitivity analyses

**a)** Adding relevant interaction terms (e.g., sex by length of stay) to the multivariable regression did not result in an increased AUROC (not shown). **b)** In the main analysis above, we combined episodes with *any* opioid, by cumulating available categories (*heroin*, *illicit methadone*, and *other opiates and synthetics*). In a sensitivity analysis, we examined only episodes with *heroin* as the primary substance. The sex disparity in outcomes was also observable in this subset (not shown). **c)** Polysubstance use is an important feature of opioid use disorder(Butelman et al., 2019; Krawczyk et al., 2021; Luo et al., 2023). As a follow-up, we examined the subset with an opioid as primary substance *and* either cocaine or methamphetamine as secondary substance. In a univariate analysis, this subset also exhibited a similar sex disparity (not shown).

## 4 Discussion

This is one of the first nationally representative “real world” studies on non-medical opioid use (considered an undesirable outcome) prior to discharge from MAT. Overall, males had greater adjusted odds of use, compared to females. More broadly, this study also identified socio-demographic and clinical conditions associated with this greater odds of this undesirable outcome, and specific conditions under which this sex disparity occurred.

### 4.1 Length of stay

Shorter lengths of stay in treatment overall (i.e., analyzing males and females together) resulted in greater odds of NMOU. This is consistent with studies showing that longer duration of MAT is associated with other improved outcomes (Kimber et al., 2010; Wakeman et al., 2020) (e.g. decreased overdose incidence). Intriguingly, a sex difference emerged at longer treatment durations (i.e., ≥366 days), where males had greater odds of NMOU compared to females (Krawczyk et al., 2021; Wakeman et al., 2020; Williams et al., 2018).

### 4.2 Route of non-medical opioid administration

Overall, persons for whom the main route of use was by inhalation (i.e., snorting), smoking (e.g., “chasing the dragon”) or injection had higher odds of NMOU compared to those using by the oral route. Increased frequency of opioid use by inhalation and smoking routes has recently been reported (Fischer et al., 2023; Huhn et al., 2018; Tanz et al., 2024), in parallel with increases in use of potent synthetic opioids (Ciccarone, 2017; Palamar et al., 2022; Rosenblum et al., 2020). Inhaled opioids cause less potent effects than after i.v. injection, likely due to pharmacokinetic mechanisms (Comer et al., 1999; Foster et al., 2008; Jones et al., 2014; Lofwall et al., 2012); less information is available for the smoking route (Hendriks et al., 2001; Jenkins et al., 1994). Importantly, these data show that inhalation (and smoking) result in outcomes comparable with injection. A sex disparity was observed in the inhalation route, in which males had greater odds of NMOU, compared to females. Overall, opioid use by inhalation and smoking routes (and relevant sex disparities) need to be investigated, given the relatively negative outcomes observed here.

### 4.3 Age at admission

Overall, persons admitted to MAT in the age bin 40-49 had greater odds of NMOU, compared to the reference category (age ≥50). While some studies report that older patients in MAT have better outcomes (Carew and Comiskey, 2018; Rajaratnam et al., 2009), this study focused specifically on *first* episodes of treatment, and it is possible that persons who enter their *first* MAT in middle age have sub-optimal outcomes. Focusing specifically on sex disparities across the lifespan, males had greater odds of NMOU compared to females, at younger age categories (18-29 and 30-39).

### 4.4 Social determinants

Social determinants, including housing and employment (as well as race and ethnicity) can affect outcomes of MAT, as well as other health interventions (Braveman and Gottlieb, 2014). Unhoused persons had greater odds of NMOU, compared to those in independent housing. Conversely, persons in “dependent” (e.g., supportive) housing had decreased odds of NMOU, compared to those in independent housing. Being unhoused is associated with diverse biopsychosocial stressors and difficulty in treatment adherence, which could underlie sub-optimal MAT outcomes (Fine et al., 2024; Gaeta Gazzola et al., 2023). Conversely, dependent housing could result in improved MAT outcomes, potentially due to biopsychosocial “wraparound” support (McLaughlin et al., 2021; Miller-Archie et al., 2019). Intriguingly, a sex disparity (greater NMOU in males vs females) was observed only in persons in independent housing, not those who are unhoused or in dependent housing.

Persons who were not in the labor force, were unemployed, or who worked part-time, had greater odds of NMOU, compared to those who were employed full-time, consistent with disparities based on economic disadvantage (Baird et al., 2022; Gaeta Gazzola et al., 2023; Han et al., 2022). Further “real world” research is needed to explore potential contributing factors within these groups, and related sex/gender differences.

#### 4.4.1 Racial background and ethnicity

African-American/black persons had greater odds of NMOU, compared to white persons. Also, hispanic/latinos had greater odds, compared to non-hispanic/latinos. Socioeconomic segregation can affect healthcare resources including MAT (Barnett et al., 2023), and racial/ethnic factors may affect the likelihood of being prescribed a specific medication (e.g., methadone versus buprenorphine) (Goedel et al., 2020; Hansen et al., 2013).

### 4.5 Psychiatric comorbidities

Psychiatric comorbidities (e.g., depression, anxiety, PTSD) are relatively frequent in individuals with opioid use disorders (Butelman et al., 2017; Gelkopf et al., 2006), and there may be bidirectional causal links between psychiatric comorbidities and the trajectory of opioid use disorder (Martins et al., 2009; Rogers et al., 2021; Rosoff et al., 2020). Males in MAT had lower odds of psychiatric comorbidity compared to females, consistent with prior studies (Braciszewski et al., 2022; Huhn et al., 2019). Prior studies show complex findings on whether psychiatric comorbidities are associated with differential MAT outcomes, based on methodological differences (Choi et al., 2015; Gelkopf et al., 2006; Krawczyk et al., 2017; Rosic et al., 2017; Zhu et al., 2021). In this nationally representative sample, presence of a psychiatric comorbidity was associated with *decreased* odds of NMOU. It may be hypothesized that being in MAT is associated with broader biopsychosocial support and psychiatric care (Hammond et al., 2022), supporting improved outcomes (Martins et al., 2009). Mu-, kappa-, and delta-opioid receptor systems (at which methadone, buprenorphine or naltrexone have differential pharmacological profiles) are involved in depression, anxiety and stress adaptation mechanisms (Bidlack et al., 2018; Browne et al., 2018; Huang et al., 2016; Jacobson et al., 2020; Perrine et al., 2006). Future large-scale studies should examine whether specific comorbid diagnoses are associated with the outcomes for specific medications, in a sex-related manner (Huhn et al., 2019).

### 4.6 Court or criminal justice referral

Persons with a court/criminal justice referral had decreased adjusted odds of NMOU, compared the larger group without such a referral. Some prior studies indicate that persons with court/criminal justice referrals have relatively improved clinical outcomes of MAT (Coviello et al., 2013; Lucabeche and Quinn, 2022; Stahler et al., 2022). Intriguingly, there was no sex disparity in outcome among those with a court/criminal justice referral, while there was a disparity (greater odds of use in males vs females) in those referred to treatment through other means (self-referral, medical, etc).

### 4.7 Geographic disparities

We adjusted for census divisions, to account for major environmental variables (Conway et al., 2023; Grimm, 2020). As shown before, there were geographic differences in overall numbers of episodes, and outcomes of MAT (Table 1 and S1-S2) (Krawczyk et al., 2021; Stahler and Mennis, 2018).

### 4.8 Limitations

Some limitations of the study should be considered: **a)** The “gender” variable in TEDS-D is coded as male/female (i.e., typical definitions of sex); further real world investigation of gender on MAT outcomes is needed (Bahji et al., 2023; Paschen-Wolff et al., 2023). **b)** There is potential selection bias, based on publicly-funded clinics in TEDS-D, compared to other MAT settings (Xu et al., 2024). **c)** Overall, cross-sectional studies cannot be used to clearly discern causality, although features such as sex, racial background and ethnicity can be linked to long-term health-related consequences, potentially through social determinants.

### 4.9 Conclusions and future studies

Males had a 1.19 adjusted odds ratio of an unfavorable outcome (NMOU) in their first MAT episode, compared to females in this real world study. Recent nationally representative data also show that heroin and synthetic opioid overdose mortality rates in males are approximately 2.5-fold greater than in females, even adjusting for overall sex-specific NMOU (Butelman et al., 2023). Overall, it may be hypothesized that several mechanisms, including MAT outcomes and riskier patterns of NMOU (e.g., higher doses, using alone) could underlie sex disparities in opioid-induced morbidity and mortality (Gicquelais et al., 2022; Kariisa et al., 2019).

While females can have specific disadvantages in MAT access (Marsh et al., 2021), this study shows that once in treatment, their outcome can be relatively more favorable than that of males. Of note, females who are unhoused, have psychiatric comorbidity or have a legal/criminal referral have similar outcomes to males in those conditions. Therefore, these conditions can put females at greater relative risk for negative MAT outcomes. Strengthening biopsychosocial support in females with opioid use disorder, and transitioning from court/criminal settings into MAT, are therefore important intervention targets (Kreek et al., 2019; Strang et al., 2020; Wakeman et al., 2020). More broadly, large scale studies should examine multiple potential mechanisms for the observed disparity in MAT outcomes, such as differences in risk-taking behaviors and executive functions (de Wit, 2009; Weidacker et al., 2023), and gender-based stigma or social/family buffering (Jalali et al., 2020; Polenick et al., 2022; Smith et al., 2021; Williams and Latkin, 2007).

## Supporting information

STROBE Checklist

Study Flowchart

Supplementary Results

## Data Availability

All raw data are available from the originator:
SAMHSA

https://www.datafiles.samhsa.gov/dataset/teds-d-2019-ds0001-teds-d-2019-ds0001

## Abbreviations

95%CI: 95% confidence intervals
AUROC: Area under the receiver-operating curve
df: degrees of freedom
LOS: Length of service (duration of treatment episode)
MAT: medication-assisted therapy for opioid use disorders
NMOU: non-medical opioid use in the month prior to discharge (binary outcome under study)
NS: Not significant
Ref: Reference category for multivariable logistic regression
TEDS-D: Treatment episode data set - Discharges
MOR: mu-opioid receptor

## Author Disclosures

### Contributors

Conceptualization, design and formal analysis (ERB, YH, RZG, NAK); data interpretation, writing and editing (ERB, YH, AM, CS, RZG, NDV, NAK).

## Acknowledgements

This work was supported by NIDA U01DA053625 (ERB), and NIDA 1RO1DA048301-01A1 (RZG), and NIDA 1RO1DA049547 (NAK), as well as NIDA Intramural funds, and from Samaritan Daytop Village.

## Declaration of generative AI in scientific writing

The authors have not used generative AI for scientific writing.

## Submission declaration and verification

These data have not been published or deposited in pre-print form.

