## Supplementary material for "Sex disparities in outcome of medication-assisted therapy of opioid use disorder: Nationally representative study": Study Flowchart

| **Full data set: TEDS-D 2019 (n=1,722,503 episodes)** | | | |  |
| --- | --- | --- | --- | --- |
| **Variable** | **Variable name** | **Filter codes** | **n=remaining** | **n=missing (excluded)** |
| Opioids as primary substance at admission | SUB1 | 5,6,7  (heroin, non-medical methadone, other opiates and synthetics) | 521,618 | 99,426 |
| Opioids as primary substance at discharge | SUB1_D | 5,6,7  (heroin, non-medical methadone, other opiates and synthetics) | 432,118 | 44,786 |
| Medication assisted opioid therapy | METHUSE | =1 (yes) | 156,145 | 15,800 |
| Number of prior treatment episodes | NOPRIOR | =0 (no prior treatment episodes for the person) | 45,134 | 3,152 |
| Service received | SERVICE | =6,7  (Ambulatory service: intensive or non-intensive) | 41,505 | 0 (none) |
| Length of service  (>30 days selected) | LOS | =31 (31-45 days)  =32 (46-60 days)  =33 (61-90 days)  =34 (91-120 days)  =35 (121-180 days)  =36 (181-365 days)  =37 (≥366 days) | 24,321 | 0 (none) |
| Source of referral to treatment | PSOURCE | Recoded as a binary variable:  =7 (any court or criminal justice referral)  =1-6 (combined category for all non- court/criminal referrals) | 24,104 | 217 |
| Co-occurring mental and substance use disorders | PSYPROB | =1 (Yes, has comorbidity)  =2 (no comorbidity) | 22467 | 1,637 |
| Age at admission | AGE | (exclude =1,2; ages 12-17)  =3,4,5 (ages 18-29)  =6,7 (ages 30-39)  =8,9 (ages 40-49)  =10,11,12 (all ages ≥50) | 22,456 | 0 (none) |
| Gender | GENDER | =1 (Male)  =2 (Female) | 22,446 | 10 |
| Living arrangements at discharge | LIVARAG_D | =1 Homeless  =2 Dependent living (e.g., supportive housing)  =3 Independent living | 15,650 | 6,796 |
| Employment at discharge | EMPLOY_D | =1 (Employed full-time)  =2 (Employed part-time)  =3 (Unemployed)  =4 (Not in labor force) | 15,387 | 263 |
| Route of non-medical opioid administration | ROUTE1 | =1 oral  =2 smoking  =3 inhalation  =4 injection  (exclude=5; “other”) | 15,186 | 41 |
| Racial background | RACE | Recoded in 3 categories:  =4 (Black/African-American)  =1-3 and 6-9, combined into “Other”  =5 (White) | 15,013 | 173 |
| Ethnicity | ETHNIC | Recoded as a binary variable:  =1-3, 5 (All Hispanic/latino origins combined)  =4 (not of Hispanic or latino origin) | 14,871 | 142 |
| Census division | DIVISION | =1 (New England)  =2 (Middle Atlantic)  =3 (East North Central)  =4 (West North Central)  =5 (South Atlantic)  =6 (East South Central)  =7 (West South Central)  =8 (Mountain)  =9 (Pacific) | 14,828 | 0 (none) |
| Reason for end of episode | REASON | Exclude=5,6 (incarcerated,  deceased) | 14,359 | 0 (none) |
| **Outcome:**  Non-medical opioid use (NMOU) in the month prior to discharge | FREQ1_D | Recoded as a binary variable:  =1 (no use)  =2,3 (some use, daily use) | **11,549** | 2,810 |
