## Supplementary Results for "Sex disparities in outcome of medication-assisted therapy of opioid use disorder: Nationally representative study"

**Table S1. Demographic variables of the analyzed sample, stratified by sex**

| **Variable** | **Condition** | **Male:**  **total n=6,685** | **Female:**  **total n=4,864** | **Contingency analysis** |
| --- | --- | --- | --- | --- |
|  |  | **N (%)** | **N (%)** |  |
| **Length of stay: (days)** | **31-45** | 788 (11.8) | 564 (11.6) | **Chi^2^ [6]=11.92; NS** |
|  | **46-60** | 541 (8.1) | 377 (7.8) |  |
|  | **61-90** | 775 (11.6) | 556 (11.4) |  |
|  | **91-120** | 634 (9.5) | 417 (8.6) |  |
|  | **121-180** | 914 (13.7) | 618 (12.7) |  |
|  | **181-365** | 1367 (20.4) | 998 (20.5) |  |
|  | **≥366*** | 1666 (24.9) | 1334 (27.4) |  |
| **Route of non-medical opioid use** | **Oral*** | 1253 (18.7) | 1259 (26.9) | **Chi^2^ [3]=99.53; p<0.0001** |
|  | **Smoking** | 473 (7.1) | 329 (6.8) |  |
|  | **Inhalation** | 2166 (32.4) | 1290 (26.5) |  |
|  | **Injection** | 2793 (41.8) | 1986 (40.9) |  |
| **Age (years)** | **18-29** | 1606 (24.0) | 1394 (28.7) | **Chi^2^ [3]=88.56 p<0.0001** |
|  | **30-39** | 2323 (34.7) | 1831 (37.6) |  |
|  | **40-49** | 1268 (19.0) | 862 (17.7) |  |
|  | **≥50*** | 1488 (22.3) | 777 (16.0) |  |
| **Housing at Discharge** | **Unhoused** | 708 (10.6) | 420 (8.6) | **Chi^2^ [2]=12.30 p=0.0021** |
|  | **Dependent living** | 1041 (15.6) | 783 (16.1) |  |
|  | **Independent living*** | 4936 (73.8) | 3661 (75.3) |  |
| **Employment at Discharge** | **Full time* employed** | 1727 (25.8) | 739 (15.2) | **Chi^2^ [3]=197.0;p<0.0001** |
|  | **Part-time employed** | 671 (10.0) | 554 (11.4) |  |
|  | **Unemployed** | 2280 (34.1) | 1792 (36.8) |  |
|  | **Not in labor force** | 2007 (30.0) | 1779 (36.6) |  |

| **Table S1 continued** | | **Male**  **total n=6,685** | **Female**  **total n=4,864** | **Contingency analysis** |
| --- | --- | --- | --- | --- |
|  |  | **N (%)** | **N (%)** |  |
| **Racial background** | **African- American/ Black** | 1029 (15.4) | 568 (11.7) | **Chi^2^ [2]=64.20; p<0.0001** |
|  | **White*** | 4714 (70.5) | 3754 (77.2) |  |
|  | **Other** | 942 (14.1) | 542 (11.1) |  |
| **Ethnicity** | **Hispanic/latino** | 945 (14.1) | 510 (10.5) | **Chi^2^ [1]=34.08; p<0.0001** |
|  | **Non-hispanic/**  **latino*** | 5740 (85.9) | 4354 (89.5) |  |
| **Psychiatric**  **comorbidity** | **Yes** | 2150 (32.2) | 2142 (44.0) | **Chi^2^ [1]=170.10; p<0.0001** |
|  | **No*** | 4535 (67.8) | 2722 (56.0) |  |
| **Court or criminal justice referral** | **Yes** | 376 (5.6) | 244 (5.0) | **Chi^2^ [1]=2.05; NS** |
|  | **No*** | 6309 (94.4) | 4620 (95.0) |  |
| **Census Division** | **New England** | 915 (13.7) | 587 (12.1) | **Chi^2^ [8]=77.64; p<0.0001** |
|  | **Middle Atlantic*** | 1589 (23.8) | 1018 (20.9) |  |
|  | **East North Central** | 696 (10.4) | 658 (13.5) |  |
|  | **West North Central** | 56 (1**) | 51 (1**) |  |
|  | **South Atlantic** | 1369 (20.5) | 954 (19.6) |  |
|  | **East South Central** | 230 (3.4) | 255 (5.2) |  |
|  | **West South Central** | 45 (1**) | 35 (1**) |  |
|  | **Mountain** | 220 (3.3) | 224 (4.6) |  |
|  | **Pacific** | 1565 (23.4) | 1082 (22.2) |  |

****reference category used in multivariable logistic regression (see Table S2).***

****rounded up to 1% due to n<100**

**Table S2**: Multivariable logistic regression for non-medical opioid use (NMOU) in the month prior to discharge from MAT (versus no use; binary variable)

| **Variable** | | **Beta coefficient** | | **Odds ratio** | |  |
| --- | --- | --- | --- | --- | --- | --- |
|  |  | **Estimate** | **95% CI** | **Estimate** | **95% CI** | **p value** |
| **Intercept** | | -0.84 | -1.04 to -0.64 | 0.43 | 0.36 to 0.53 | N/A |
| **Sex**  Ref: Female | Male | 0.17 | 0.086 to 0.26 | 1.19 | 1.09 to 1.29 | <0.0001 |
| **Length of Stay**  Reference category (Ref):  ≥366 days | 31-45 days | 1.15 | 1.00 to 1.31 | 3.16 | 2.71 to 3.70 | <0.0001 |
|  | 46-60 days | 1.06 | 0.89 to 1.24 | 2.88 | 2.42 to 3.44 | <0.0001 |
|  | 61-90 days | 0.82 | 0.67 to 0.96 | 2.26 | 1.95 to 2.62 | <0.0001 |
|  | 91-120 days | 0.78 | 0.62 to 0.94 | 2.18 | 1.86 to 2.56 | <0.0001 |
|  | 121-180 days | 0.55 | 0.42 to 0.69 | 1.74 | 1.52 to 2.00 | <0.0001 |
|  | 181-365 days | 0.44 | 0.32 to 0.56 | 1.55 | 1.38 to 1.74 | <0.0001 |
| **Route of non-medical opioid use**  Ref: Oral | Injection | 0.58 | 0.47 to 0.69 | 1.79 | 1.60 to 2.00 | <0.0001 |
|  | Inhalation | 0.52 | 0.40 to 0.64 | 1.69 | 1.50 to 1.90 | <0.0001 |
|  | Smoking | 0.74 | 0.54 to 0.94 | 2.09 | 1.72 to 2.55 | <0.0001 |
| **Age**  **Ref: ≥50** | 40-49 | 0.22 | 0.079 to 0.36 | 1.24 | 1.08 to 1.43 | 0.0021 |
|  | 30-39 | 0.01 | -0.11 to 0.14 | 1.02 | 0.90 to 1.15 | NS |
|  | 19-29 | -0.03 | -0.16 to 0.10 | 0.97 | 0.85 to 1.11 | NS |
| **Housing at discharge**  Ref: Independent living | Unhoused | 0.31 | 0.15 to 0.46 | 1.36 | 1.164 to 1.59 | 0.0001 |
|  | Dependent living | -0.56 | -0.68 to -0.44 | 0.57 | 0.51 to 0.64 | <0.0001 |
| **Employment at discharge**  Ref: Full-time employed | Not in labor force | 0.42 | 0.30 to 0.54 | 1.52 | 1.35 to 1.72 | <0.0001 |
|  | Unemployed | 0.54 | 0.42 to 0.66 | 1.72 | 1.53 to 1.93 | <0.0001 |
|  | Part-time  employed | 0.21 | 0.062 to 0.36 | 1.24 | 1.06 to 1.44 | 0.0057 |
| **Race**  Ref: White | “Other” | -0.13 | -0.28 to 0.028 | 0.88 | 0.76 to 1.03 | NS |
|  | African-American / Black | 0.73 | 0.59 to 0.87 | 2.07 | 1.80 to 2.39 | <0.0001 |
| **Ethnicity**  Ref: Not Hispanic / Latino | Hispanic or Latino | 0.32 | 0.17 to 0.48 | 1.38 | 1.180 to 1.62 | <0.0001 |
| **Psychiatric Comorbidity**  Ref: no comorbidity | Comorbidity present | -0.26 | -0.35 to -0.17 | 0.77 | 0.71 to 0.85 | <0.0001 |
| **Court or criminal justice referral**  Ref: non- court /criminal referral | Court/criminal referral | -1.15 | -1.34 to -0.96 | 0.32 | 0.26 to 0.38 | <0.0001 |
| **Census Division**  Ref: Middle Atlantic | New England | -0.97 | -1.12 to -0.83 | 0.38 | 0.33 to 0.44 | <0.0001 |
|  | East North Central | -0.65 | -0.80 to -0.50 | 0.52 | 0.45 to 0.61 | <0.0001 |
|  | West North Central | -0.20 | -0.61 to 0.23 | 0.83 | 0.54 to 1.26 | NS |
|  | South Atlantic | 0.43 | 0.29 to 0.57 | 1.53 | 1.33 to 1.76 | <0.0001 |
|  | East South Central | -0.23 | -0.44 to -0.014 | 0.80 | 0.64 to 0.98 | 0.04 |
|  | West South Central | 0.41 | -0.069 to 0.92 | 1.51 | 0.93 to 2.51 | NS |
|  | Mountain | -0.80 | -1.029 to -0.57 | 0.45 | 0.35 to 0.57 | <0.0001 |
|  | Pacific | 0.20 | 0.056 to 0.33 | 1.22 | 1.057 to 1.40 | 0.006 |

***Ref:** Reference category
